# Factors affecting social integration after road traffic orthopaedic injuries in Rwanda

**DOI:** 10.1101/2023.08.03.23293597

**Authors:** JC Allen Ingabire, Aimee Stewart, Carine Uwakunda, Didace Mugisha, JB Sagahutu, Gerard Urimubenshi, David K. Tumusiime, Georges Bucyibaruta

**Affiliations:** Department of Surgery, University Teaching Hospital of Kigali, University of Rwanda, Rwanda; Physiotherapy Department, University of the Witwatersrand, South Africa; Department of Surgery, Kibagabaga Level II Teaching Hospital, Rwanda; Department of Environmental, University of Rwanda, Rwanda; Physiotherapy Department, University of Rwanda, Rwanda; Department of Epidemiology and Biostatistics, Imperial College London, UK

**Author notes:** Correspondence:JC Allen Ingabire.

**Keywords:** Social integration, road traffic orthopaedic injuries, activities and participation, IMPACT-S, rehabilitation

## Abstract

**Background:** Road traffic injuries (RTIs) leading to long-term disability present a significant public health challenge, causing immense personal and societal consequences. However, in many developing countries, information on the social integration of patients post-RTI remains limited.

**Purpose:** This study aimed to identify factors contributing to social integration following road traffic-related orthopedic injuries in Rwanda.

**Methodology:** The research encompassed a multicenter, cross-sectional study involving 369 adult RTI victims from five Rwandan referral hospitals, all of whom experienced accidents in 2019. Participants completed the IMPACT-S Questionnaire, which evaluated the level of social integration in terms of activities and participation. Statistical analysis using logistic regression, with a significance level set at p<0.05, helped estimate odds ratios (OR) and 95% confidence intervals (CI). We obtained ethical approval to conduct the study from the University of Rwanda, College of Medicine and Health Sciences Institutional Review Board. All participants signed a written consent before enrollment into the study, and all data were kept confidential and only used for the purpose of this study.

**Results:** The study’s findings indicated that the mean age of RTI victims was 37.5±11.26 years, with a notable male predominance over females. Of the participants, 5.69% were unable to resume normal life activities. The overall mean score on the IMPACT-S scale was moderate, at 77±17. Specifically, participants achieved an average score of 76±16 for “activities” and a higher average of 84±16 for “participation.” Certain factors were associated with poor social integration compared to others, including belonging to the age group above 65 years (OR=8.25, p=0.02), female sex (OR=3.26, p=0.02), lack of rehabilitation (OR=3.82, p=0.01), and length of hospital stay > 15 days (OR=4.44, p=0.02).

**Conclusion:** The majority of RTI victims in Rwanda achieved successful reintegration into society; nevertheless, their mobility and community engagement were more significantly impacted compared to other aspects assessed by the IMPACT-S scale. The study emphasized the importance of early management, effective rehabilitation, and prompt patient discharge from the hospital in facilitating a successful return to everyday life after road traffic-related orthopedic injuries.

## Background

Long-term disability resulting from Road Traffic Injuries (RTIs) is a pressing public health concern with devastating effects on individuals and significant societal and economic impacts worldwide [1], [2]. Annually, around 50 million people suffer injuries and 1.2 million lose their lives due to road traffic accidents, leaving 30% of survivors with permanent disabilities and 14% unable to return to work[3]–[5]. This primarily affects the working-age population in low and middle-income countries (LMICs), creating profound consequences for individuals, society, and the economy[6].

Effective management of injured patients aims to restore their normal functioning, and various biopsychosocial factors influence post-RTI functional outcomes[7], [8]. Social integration of patients post-RTI is a key outcome of successful management, and early psychological support and educating family members play vital roles in promoting social reintegration [6], [9][10].

The International Classification of Functioning, Disability, and Health (ICF) defines participation in life as a crucial health outcome, encompassing an individual’s involvement in society’s usual activities[11]. Social integration, as defined by the ICF, necessitates interventions to facilitate interaction with the environment for optimal performance in an individual’s life[12]–[14]. Various instruments, such as the IMPACT-S questionnaire, measure participation and activities following the ICF guidelines[15].

However, individuals with disabilities may encounter challenges in acceptance by their families, limited job opportunities, and difficulties in reintegrating into society[16]–[18]. Adequate rehabilitative care is essential for positive functional outcomes and social reintegration, particularly in LMICs[19]. Rwanda faces a significant number of RTI victims[20], but the limited rehabilitation centers and personnel hinder their social reintegration[21].

This study employs the IMPACT-S questionnaire to identify factors contributing to social integration after road traffic orthopedic injuries in Rwanda, aiming to shed light on improving outcomes and addressing the challenges faced by RTI victims in the country.

## Methodology

### Study design and study settings

A multi-centre cross-sectional study was undertaken to analyze hospital-based data on road traffic-related orthopaedic injuries that occurred in 2019 and were treated at the five Rwandan referral hospitals. These hospitals are referral and teaching hospitals with emergency, orthopaedic, mental health departments, and rehabilitation services. The study took place from the 1^st^ March 2022 to 31^st^ August 2022, two years after the injuries occurred, at Centre Hospitalier Universitaire de Kigali (CHUK), Rwanda Military Hospital (RMH), and King Faisal Hospital (KFH), all located in Kigali City, but which receive patients from across Rwanda. The other two hospitals are Centre Hospitalier Universitaire (CHUB) in the Southern Province and Ruhengeri Hospital (RH) in the Northern Province.

### Study population and sample size

The study population comprised 2019 road traffic injury (RTI) survivors aged 18 and above admitted to the above five hospitals for both upper and lower limbs injuries. According to the records of the above five mentioned hospitals, around 4,600 cases post-RTIs with 1986 orthopaedic injuries were admitted during the selected study period. We used Krejcie and Morgan’s formula[22] for sample calculation and random sampling for sample size. The sample size representative of these RTI victims was 369.

We consulted the hospital records from the emergency departments, outpatients and admission for patients’ demographics and contacts, details of the injury pattern, and the length of stay in the hospital. We excluded participants who were not oriented to time and space and could not respond to the questionnaire and patients with injuries other than orthopaedic. Those fulfilling the inclusion criteria of being above 18 years and having an orthopaedic road injury in 2019 were contacted via telephone for their demographic details and requested to come to the hospital for further evaluation.

### Psychometric properties of the instruments

Participation and activities (Social integration) were evaluated using IMPACT-S (ICF Measure of Participation and ACTivities), an ICF-based participation tool called Patient Reported Outcome Measures (PROMS). The measure is designed to describe functioning and disability independent of health conditions and guide the participation level of patients with disabilities. This tool consists of 32 items grouped into nine domains (learning and applying knowledge, general tasks and demands, communication, mobility, self-care, domestic life, interpersonal interactions and relationships, major life areas, community, social and civic life). The IMPACT-S also has two sub-total scores for Activities and Participation and one IMPACT-S total score. All summary scores were averaged item scores, converted into 0–100 scales. Higher IMPACT-S scores reflect better functioning (or less disability), meaning nearly entirely socially reintegrated in life after a road traffic injury.

This tool was validated by Marcel Post et al., 2008, in 197 road traffic survivors[23] with good psychometric properties according to the ICF framework. The IMPACT-S tool has been validated in conditions like carpal tunnel syndrome[24] and many languages, including Turkish[25]. Amir Javanmard et al.2020 compared six instruments used in the participation and activities evaluation for patients with spinal cord injuries and found that the IMPACT-Shas had higher psychometric measures than other instruments [26]. The questionnaire was translated from English to Kinyarwanda by two language experts and back to English by two other language experts to address the cultural and linguistic equivalence, and the responses were the same. Also, we sent the questionnaire to experts in orthopaedic and rehabilitation for their opinion on the quality of translation, clarity and suitability for the Rwandan participants.

### Procedure

Of the 1986 patients with orthopaedic injuries, we contacted 1721 on the phone; some had died, or their phones malfunctioned. The severity of the injury was evaluated using the Kampala Trauma Score (KTS), which is classified as mild, moderate and severe. After sampling, participants were invited to the hospital to assess their current status after almost two years post-RTIs. Using the IMPACT-S questionnaire, we measured the patient’s overall level of social integration (participation and activities) after road traffic orthopaedic injuries in Rwanda. Participants filled out the questionnaire by considering how much their impairments interfered with their lives in the last 30 days before the interview. They answered on a 4-point response scale from 0-3 (Extreme, considerable, some and no limitations), and the research assistants helped the participants to complete the questionnaire if they could not write.

We calculated each IMPACT-S domain’s mean and standard deviation (learning and applying knowledge, general tasks and demands, communication, mobility, self-care, domestic life, interpersonal interactions and relationships, major life areas, community, social and civic life).

The participant’s socioeconomic status (Ubudehe) was collected according to the Rwanda government classification, where category I include impoverished and vulnerable citizens. Category II includes citizens who can afford some form of rented or owned accommodation but are not gainfully employed and can only afford to eat once or twice a day. Category III includes citizens who were gainfully employed or employers of labour. Category IV are citizens who are chief executive officers of big businesses, full-time employees with organisations, industries or companies, government employees, owners of shops or markets and owners of commercial transport vehicles or trucks [27].

The study’s primary outcome is social integration (activities and participation). The risk factors include demographic data, the Kampala Trauma Scale, length of hospital stay, and rehabilitation.

### Data management and Statistical analysis

Data were collected using the questionnaires, entered into a computer by a Google form data entry mode, and analysed using the R Software. We performed a descriptive analysis of the patient-reported outcome measure scale (IMPACT-S). Categorical variables were summarised using frequencies and percentages, continuous variables with means and standard deviations (SD). We used a student’s t-test to compare continuous variables and the Chi-Square test for nominal (categorical) variables. We utilised multivariate logistic regression to assess associations between risk factors and IMPACT-S score categories. We considered the *P-value* < 0.05 to be statistically significant.

### Ethical consideration

We obtained ethical approval to conduct the study from the University of Rwanda, College of Medicine and Health Sciences Institutional Review Board (18/CMHS IRB/2022). The Rwanda National Research Committee operating in the Ministry of Health approved this study (NHRC/2022/PROT/014), and we collaborated with the Rwanda Biomedical Center (5535/RBC/2022) injury department. We obtained local ethical approvals from the five hospitals’ ethics committees; CHUK(EC/CHUK/051/2022), CHUB (REC/UTHB/089/2022), RH(313/RRH/DG/2022), KFH(EC/KFH/015/2022), RMH(RMH IRB/027/2022). We obtained written consent from all participants before enrollment into the study, after explaining the purpose of the study, and all data were kept confidential and only used for the purpose of this study.

## Results

### Demographic characteristics of the participants

Based on the data provided in Table 1, a total of 369 individuals responded to the survey. Among these, 64.5% (238 participants) were recruited from CHUK. The average age of all participants was 37.5±11.26 years, with the majority falling within the age range of 31-50 years. Males constituted the majority at 74.25%. Approximately 41.73% (172) of all participants attended primary school, and 46.34% (171) resided in Kigali city. A significant portion, 41.73% (154), were engaged in business, while 29% (107) were part of the informal sector without fixed employment. The majority of our participants belonged to category III of the socioeconomic class (Ubudehe), comprising 61.52% (227) of individuals. This was followed by category II, which represented 33.06% of the participants. Additionally, 61.52% of the reported injuries were associated with motorcycle-related accidents.

**Table1:** Demographic characteristics of the participants.

| Factors | n(%) | Factors | n(%) |
| --- | --- | --- | --- |
| **Hospital** |  | **Level of education** |  |
| CHUK | 238(64.50) | None | 28(7.58) |
| CHUB | 32(8.67) | Primary | 172(41.73) |
| RH | 29(7.86) | Secondary | 110(29.81) |
| RMH | 30(8.13) | University | 59(15.99) |
| KFH | 40(10.84) | **Residence** |  |
| **Age group** | Mean= 37.57(±11.26) | Kigali City | 171(46.34) |
| 18-30 | 102(27.64) | Secondary cities | 94(25.47) |
| 31-45 | 199(53.93) | Other Districts | 104(28.18) |
| 46-65 | 59(15.99) | **Occupation** |  |
| >65 | 9(2.44) | Farmer | 31(8.40) |
| **Sex** |  | Business | 154(41.73) |
| Male | 274(74.25) | Students | 5(1.36) |
| Female | 95(25.75) | Public service | 58(15.72) |
| **Marital status** |  | Informal sector | 107(29.00) |
| Single | 87(23.58) | Retired | 14(3.79) |
| Married | 265(71.82) | **Socio-economic status (Ubudehe)** |  |
| Divorced | 8(2.17) | I | 20(5.42) |
| Other | 9(2.44) | II | 122(33.06) |
|  |  | III | 227(61.52) |

Source: Primary data

### Clinical factors

“Table 2 shows that 52.85% (195) of all participants had isolated lower limb injuries, while polytrauma represented 21.14% (78) of cases. Half of our participants were managed within one day (49.32%), with a mean treatment duration of 30 days, and 42.01% (155) were treated with Open Reduction and Internal Fixation (ORIF). Regarding hospital stay, about 55.29% (204)were discharged within 14 days, and the mean hospital stay was 30 days. Our findings indicate that 66.84% (246) had a moderate Kampala Trauma Score (KTS). After completing their injury treatment, 37.13% of the participants were unable to undergo rehabilitation, and 5.69% experienced limitations in integrating into Rwandan society.

**Table2:** Clinical factors.

| **Factors** | **n(%)** | **Factors** | **n(%)** |
| --- | --- | --- | --- |
| **Kampala Trauma Score** |  | **Intervention** |  |
| Mild | 22(5.96) | Closed reduction+POP | 40(10.84) |
| Moderate | 247(66.94) | Open Reduction Internal Fixation | 155(42.01) |
| Severe | 100(27.1) | Open Reduction External Fixation | 57(15.45) |
| **In Hospital Diagnosis** |  | Amputation | 12(3.25) |
| Upper extremity injuries | 48(13.01) | Other | 105(28.46) |
| Lower extremity injuries | 195(52.85) | **Length of Hospital Stay** |  |
| Both upper and lower extremity injuries | 20(5.42) | 0-7 days | 149(40.38) |
| Polytrauma | 78(21.14) | 8-14 days | 55(14.91) |
| Soft tissues injuries | 28(7.59) | 15-30 days | 71(19.24) |
| **Time before management** |  | >30days | 94(25.47) |
| ≤1day | 182(49.32) | **Rehabilitation** |  |
| 2-7 days | 116(31.44) | Yes | 232(62.87) |
| 8-14 days | 23(6.23) | No | 137(37.13) |
| 15-30 days | 30(8.13) | **Level of reintegration** |  |
| >30days | 18(4.88) | No limitations(None+Some) | 348(94.30) |
|  |  | Limitations<br>(Considorable+Extreme) | 21(5.69) |

*Source: Primary data*

Table 3 shows the components of the IMPACT-S, with 32 elements of the ICF domains presented as questions. The questions were graded into four categories: extreme limitations, considerable limitations, some limitations, and no limitations in activities (18 items) and participation (14 items). Among all participants, the highest score in the “no limitations” category was 97.83% for communicating and producing, while the lowest was 15.99% for lifting and carrying objects. The category of “some limitations” ranged from 1.63% for communicating and producing to 22.49% for washing and dressing. The category of “considerable limitations ranged from 0.27% for communicating and receiving to 41.46% for community life. Extreme limitations ranged from 0.00% for communicating and producing to 55.01% for lifting and carrying objects. These findings demonstrate the impact of road traffic orthopaedic injuries on everyday life, even though most victims have fully reintegrated.

**Table 3:**
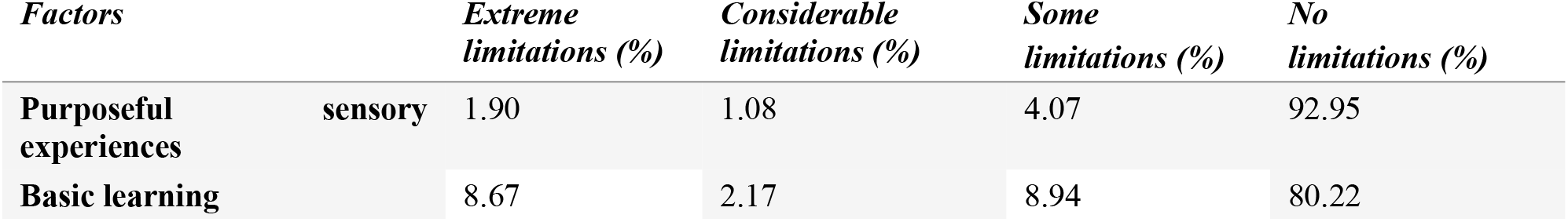
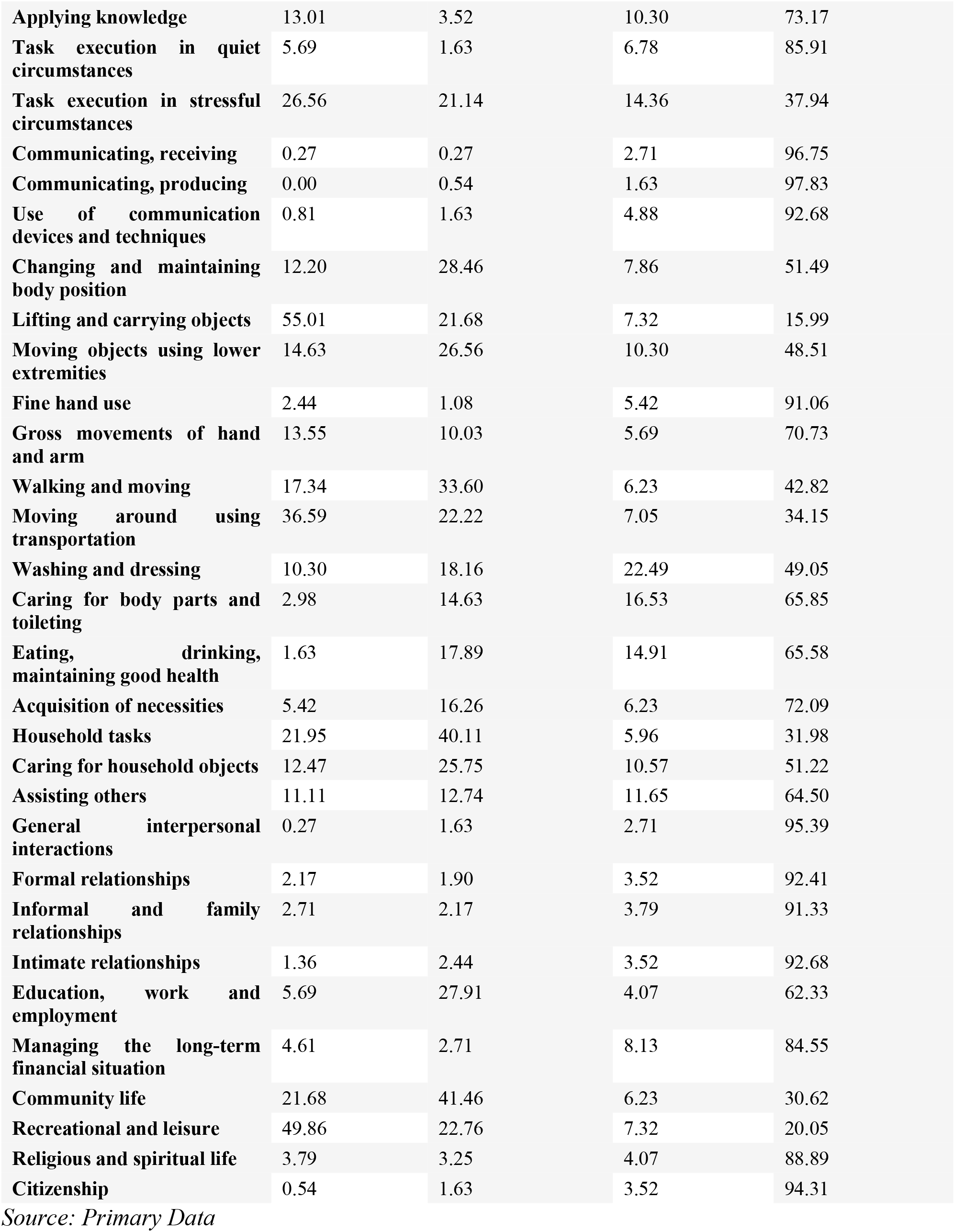
Item scores of IMPACT-S.

Based on Table 4, which summarizes the nine categories of the IMPACT-S and two subtotals (activity and participation) out of 100, the overall average score for the IMPACT-S was 77±17, indicating a favorable rating for these participants. Among the domains, participants excelled in communication, achieving the highest average score of 98±8, while mobility obtained the lowest score of 62±25. Activities had an average score of 76±16 compared to 84±16 for participation, which are the two primary components of the IMPACT-S questionnaire.

**Table 4:** IMPACT-S summary scores.

| Factors | MEAN | SD |
| --- | --- | --- |
| **Knowledge** | 88.05 | 22.92 |
| **General tasks** | 72.76 | 28.71 |
| **Communication** | 98.07 | 8.00 |
| **Mobility** | 62.27 | 24.58 |
| **Self-care** | 77.78 | 26.10 |
| **Domestic-life** | 68.59 | 29.41 |
| **Interpersonal** | 95.89 | 12.29 |
| **Major life areas** | 82.61 | 25.41 |
| **Community life** | 67.75 | 20.01 |
| **Activities total** | 76.28 | 17.81 |
| **Participation total** | 84.16 | 17.62 |
| **IMPACT-S total** | 77.10 | 16.72 |

*Source: Primary Data*

Generally, all the average scores are above 50 on a 0-100 scale, indicating that the majority of the participants have made significant progress in their recovery and can carry out and participate in most of their daily activities. Nevertheless, the average score for mobility was lower than the scores for the other domains, suggesting that some victims may still face mobility challenges due to their injuries. This highlights the importance of ongoing rehabilitation and support for the victims.

IMPACT-S scored 0 for extreme limitations, 1 for considerable limitations, 2 for some limitations, and 3 for no limitations. The binary score combined extreme and considerable limitations into one category labelled as a limitation (score 1), while the remaining categories were combined and labelled as no limitation (score 0). Poor social integration was associated with the age group >65 years (p-value<0.01) and female sex (p-value=0.04). Marital status for separated couples (p-value=0.03), people in the business category (p-value=0.01), socioeconomic status category III (p-value=0.04), lack of rehabilitation management (p-value=0.01), and length of hospital stay (p-value=0.02) were also identified as factors negatively affecting social integration (Table 5).

**Table 5:** Univariate model: Association between level of limitation and each factor.

| Factors |  | OR | CI | p-value |
| --- | --- | --- | --- | --- |
| Age group | Reference (18-30) | 0.051 | 0.01-0.11 | 0.00 |
|  | 31-45 | 0.70 | 0.22-2.44 | 0.56 |
|  | 46-65 | 1.79 | 0.48-6.72 | 0.37 |
|  | >65 | 9.7 | 1.68-50.86 | <0.01 |
| Sex | Reference (male) | 0.04 | 0.02-0.07 | 0.00 |
|  | Female | 2.50 | 0.97-6.24 | 0.04 |
| Marital Status | Reference (Single) | 0.03 | 0.00-0.09 | 0.00 |
|  | Married | 1.56 | 0.49-6.89 | 0.49 |
|  | Divorced | 4.00 | 0.18-36.43 | 0.25 |
|  | Separated | 8.00 | 0.93-56.90 | 0.03 |
| Occupation | Reference (Farmer) | 0.14 | 0.04-0.37 | 0.00 |
|  | Business | 0.13 | 0.02-0.64 | 0.01 |
|  | Student | 1.68 | 0.07-15.84 | 0.67 |
|  | Public service | 0.24 | 0.03-1.31 | 0.11 |
|  | Unemployed | 0.61 | 0.18-2.42 | 0.45 |
|  | Other | 0.51 | 0.02-3.96 | 0.57 |
| Socio-economic status | Reference (I) | 0.17 | 0.04-0.52 | 0.00 |
|  | II | 0.39 | 0.10-1.94 | 0.20 |
|  | III | 0.23 | 0.62-1.12 | 0.04 |
| Rehabilitation | Reference (Yes) | 0.03 | 0.01-0.06 | 0.00 |
|  | No | 3.36 | 1.34-9.17 | 0.01 |
| Length Hospital | Reference (1-7days) | 0.02 | 0.00-6.52 | 0.00 |
| **Stay** | 8-14days | - | - | 0.99 |
|  | 15-30days | 3.96 | 1.15-15.58 | 0.03 |
|  | >30days | 3.8 | 1.21-14.51 | 0.02 |

In the multiple logistic regression analysis, several factors were found to contribute to limitations in social reintegration after injury. The results are summarized in Table 6. Age group above 65 OR= 8.25 (95%CI =1.15-55.56), p-value=0.02.This means that individuals above the age of 65 are 8.25 times more likely to experience poor social reintegration compared to other age groups. Being female is associated with a 3.26 times higher likelihood of experiencing limitations in social reintegration with OR=3.26(95%CI =1.14-9.25), p-value=0.02.

**Table6:**
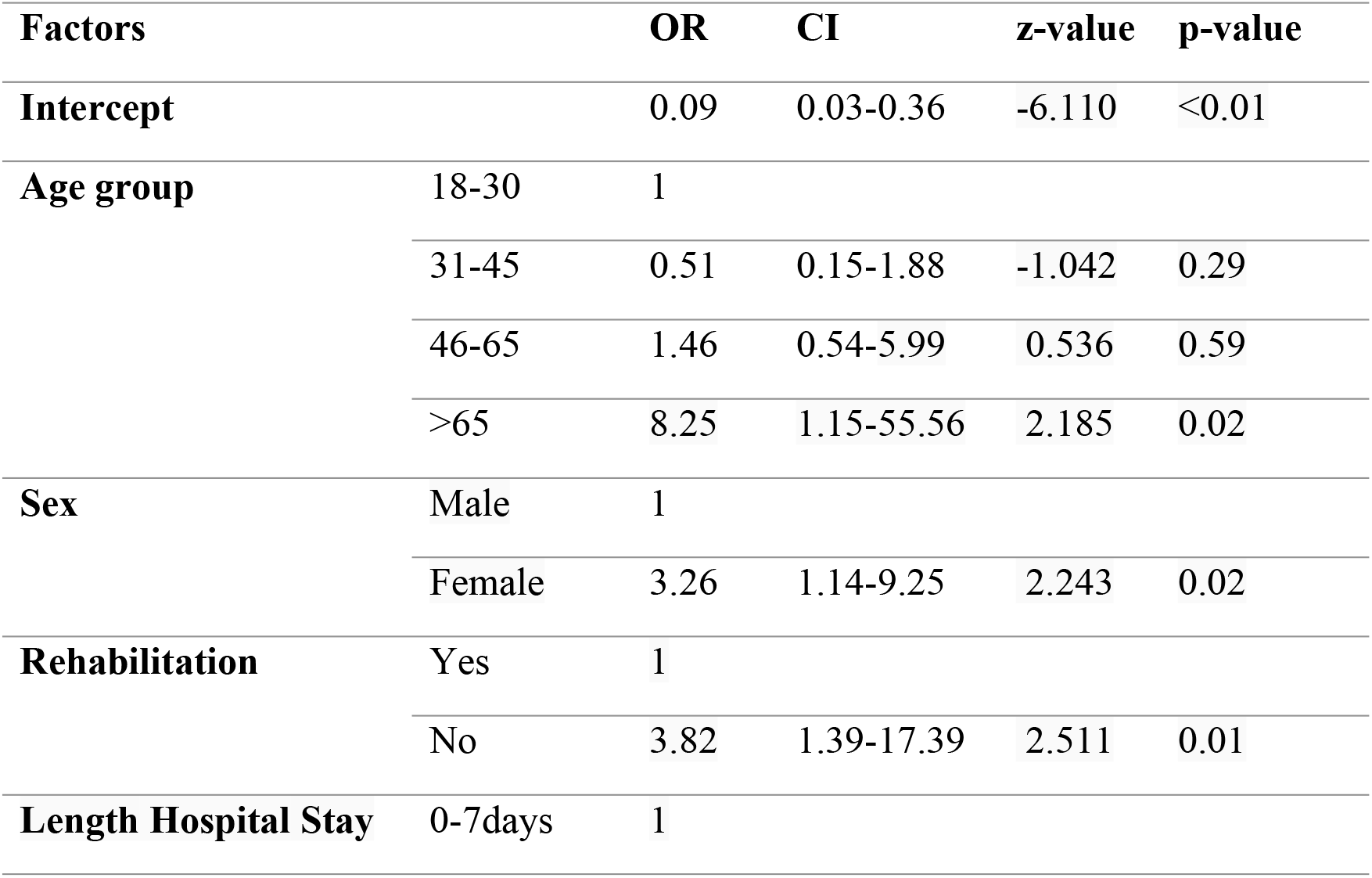
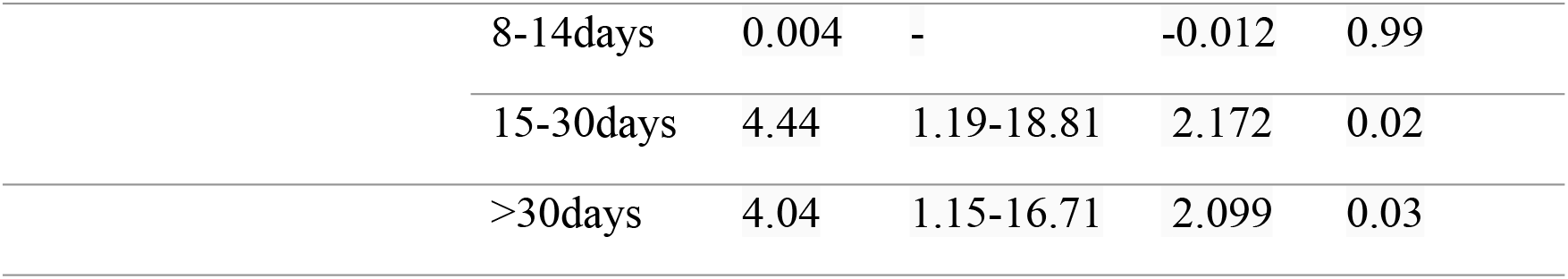
Multiple logistic regression of.

Other contributing factors to social integration limitation include lack of rehabilitation associated with a 3.82 times higher likelihood of experiencing limitations in social reintegration with OR=3.82(95%CI =1.39-17.39), p-value=0.01 and length of hospital stay of 15-30 days associated with a 4.44 times higher likelihood of experiencing limitations in social reintegration with an OR=4.44(95%CI =1.19-18.81), p-value= 0.02 as well as the length of hospital stay of >30 days with an OR=4.04(95%CI =1.15-16.71), p-value= 0.03.

## Discussion

Our study aimed to determine the level of social integration (activities and participation) following road traffic orthopaedic injuries (RTIs) in Rwanda. The findings of our study revealed several significant factors contributing to limitations in social integration after RTIs in Rwanda, including the age group above 65, female sex, lack of rehabilitation, and a hospital stay of more than two weeks.

In 2019, half of the road traffic injuries in Rwanda were limb trauma, consistent with findings from other studies conducted in LMICs[28], [29]. Males were more predominant than females, which can be explained by the higher mobility of men and their greater involvement in general activities in Rwanda, a pattern observed in other Sub-Saharan African countries as well[30][31][32]. Globally, road traffic injury victims are typically in the working age group[33]–[35] with fewer unemployed [36]–[38] and our study confirmed this finding. The mean age of our participants was 37.5 years, with a predominant representation in the age group of 31-50 years.The results of our study indicate that the majority of the RTI victims were able to integrate back into their daily activities after the accident.

More than half of the participants in our study belonged to socioeconomic class category III, which included individuals who were gainfully employed or even employers themselves. This finding highlights the association between accidents and a high rate of movement among the victims.Motorcycles were identified as the leading cause of accidents, followed by motor vehicles. As of 2021, there were over 100,000 motorcycles in Rwanda, with half of them operating as moto-taxis [39]. It is noteworthy that more than half of the victims in our study had lower limb injuries, and a quarter of them experienced polytrauma at the time of injury. This trend aligns with findings from studies conducted in LMICs, where lower limb injuries are commonly observed in road traffic injuries[40], [41].Among the orthopaedic injuries, more than half of the cases required surgical intervention, either through open reduction and internal fixation or external fixation. The average hospital stay for the participants was 30 days. It is important to note that polytrauma patients who required multiple interventions tended to have extended hospital stays.

Our findings showed that half of our participants were managed within one day (49.32%), with a mean of 30 days and 42.01% were treated by Open Reduction and Internal Fixation (ORIF). The majority were discharged within 14 days (40.38%), mean hospital stay was 30 days 246/368(66.84%) had moderate Kampala Trauma Score (KTS). After injury treatment, 37.13% of the victims could not undergo any rehabilitation management. For our study, 37% of the prescribed rehabilitation was not done after injury management, primarily due to financial issues and the long distance between their homes and the district hospitals. The same findings were observed in other studies from LMICs where access to rehabilitation ranges from 5%-59%, and in many countries, rehabilitation centres are lacking [42]–[44]. Lack of rehabilitation in post-RTI has been associated with a low rate of return to work through a significant impact on the activities and participation of the victims, which is the case in our findings. Many researchers have suggested community-based rehabilitation in post-RTI for complete social integration [45]–[47].

The primary outcome of this study was the evaluation of social integration using the Measure of Participation and Activities Screener (IMPACT-S). The results indicated that participants had higher scores in the category of no limitations for activities such as communication and production, while the lowest scores were observed in the category of lifting and carrying objects, suggesting that participants were more comfortable with communication tasks compared to tasks that required physical strength. Participants who had some limitations in activities and participation performed relatively well in communication and production but faced difficulties with tasks related to washing and dressing. The category of considerable limitations included participants who encountered significant challenges in executing community life activities, struggling with various daily tasks. More than half of the participants experienced extreme limitations when it came to lifting and carrying objects. These findings underscore the impact of road traffic orthopaedic injuries on important aspects of daily life.

The study findings revealed that while participants scored high in terms of social participation, they faced difficulties in performing activities. This can be attributed to the focus of our research on orthopaedic injuries, which predominantly affect the limbs compared to other body systems. These findings align with similar studies conducted in different countries, such as the study by M. Post et al. in 2008, which validated the IMPACT-S tool. Ahmed Nour et al. (2023) conducted a study in Cameroon and found that more than 39% of patients with limb injuries experienced difficulties with activities of daily living [23][48].These findings emphasize the need to improve rehabilitation services from the early stages of post-road traffic injuries to address the limitations in activities and promote better social integration.

Studies have consistently shown that the IMPACT-S tool is the most effective tool for summarizing all chapters of the International Classification of Functioning, Disability and Health (ICF) when compared to other tools [15], [26]. The IMPACT-S tool consists of nine domains and two subtotals.In this study, the overall IMPACT-S mean score was found to be good for the participants, which is consistent with findings reported by other authors who have also used this tool. These authors have explained that the level of activities and participation becomes acceptable after accidents [24], [25].

Among the domains of the IMPACT-S tool, communication had a higher mean score compared to mobility, which had a lower mean across all domains. This can be explained by the high number of lower limb injuries observed in this study, which is consistent with findings from other studies [23], [25]. Furthermore, the activity domain had a lower mean for the IMPACT-S subtotal compared to the participation domain. These findings can be attributed to the specific injuries sustained by these patients at the time of the accident.

After calculating the IMPACT-S scores, we analyzed the factors associated with activities and participation using a binary score. In this scoring system, scores 0 and 1 were combined to represent limitations, while scores 2 and 3 were grouped into 1 to indicate no limitations. Several factors were found to be associated with limitations in social integration. These included being above 65 years of age, female sex, being in a separated marital status, belonging to the business category for occupation, and falling into socioeconomic status category III. These findings provide insights into the univariate factors that can help explain the long-term outcomes of victims of road traffic injuries (RTIs) and their ability to return to everyday life.

Among the clinical factors, the lack of rehabilitation management was also found to contribute to limitations in social integration and longer hospital stays. These factors align with findings from other studies that have identified them as predictors of poor participation and activities in post-RTI scenarios in low- and middle-income countries (LMICs) [43], [49].

Social integration following road traffic injuries (RTIs) is a critical health outcome influenced by various factors. Through multiple logistic regression analysis, we have identified the factors that contribute to limitations in social integration among individuals post-accident. Among these factors, the age group above 65 years was found to contribute eight times more to social integration limitations compared to other age groups. Additionally, females were found to contribute three times more to these limitations compared to males. Lack of rehabilitation had a significant impact, contributing nearly four times more to limitations in social integration compared to attending rehabilitation sessions.

Furthermore, the length of hospital stay has been shown in other studies to be a determinant of social integration following RTIs [17], [40]. In our study, a hospital stay of more than two weeks contributed four times more to social integration limitations compared to individuals who spent less than two weeks in the hospital. These findings highlight the importance of considering these factors in understanding and addressing limitations in social integration among individuals recovering from RTIs.

This study will serve as a foundation for future research aimed at assessing the quality of life of individuals with long-term disabilities resulting from orthopedic injuries sustained in road traffic accidents. The findings from this study will provide valuable insights for stakeholders in developing policies and interventions to enhance activities and participation after road traffic injuries (RTIs). Consideration of rehabilitation approaches such as home-based rehabilitation or E-rehabilitation can potentially expedite the process of post-RTI social integration. These strategies can be explored further in order to improve the overall outcomes and quality of life for individuals recovering from RTIs.

By incorporating these findings into policy development, stakeholders can work towards enhancing the support and rehabilitation services available to individuals affected by long-term disabilities resulting from road traffic orthopedic injuries. This, in turn, can contribute to improving the activities and participation of individuals post-RTIs.

Our study has identified several limitations that should be acknowledged. Firstly, there was a two-year gap between the time of injury and the assessment of patient outcomes. This time lapse may have introduced variability and could affect the generalizability of our findings. Secondly, we relied on secondary data for both the baseline and follow-up measurements, which presented certain challenges. The use of existing data may have led to missing information and limited our ability to obtain a comprehensive understanding of the patients’ conditions. Furthermore, the presence of missing information in the recorded data was another limitation that impacted the generalizability of our findings. The incomplete data may have introduced biases and affected the accuracy of our analysis.

## Conclusion

Our study findings indicate that the majority of road traffic orthopedic injury victims in Rwanda are able to reintegrate into society following the accident. However, certain domains such as mobility and community life are more adversely affected than others. We identified several factors that have a negative impact on social integration after road traffic injuries in Rwanda. These factors include older age, being female, lack of rehabilitation, and longer hospital stays. The study highlights the significance of early management, rehabilitation, and timely discharge from the hospital in facilitating the return to everyday life after the accident. These factors play a crucial role in improving social integration outcomes for individuals affected by road traffic orthopedic injuries.

### Research funding

This research (or “[initials of fellow]”) was supported by the Consortium for Advanced Research Training in Africa (CARTA). CARTA is jointly led by the African Population and Health Research Center and the University of the Witwatersrand and funded by the Carnegie Corporation of New York (Grant No. G-19-57145), Sida (Grant No:**54100113**), Uppsala Monitoring Center, Norwegian Agency for Development Cooperation (Norad), and by the Wellcome Trust [reference no. 107768/Z/15/Z] and the UK Foreign, Commonwealth & Development Office, supported by the Developing Excellence in Leadership, Training and Science in Africa (DELTAS Africa) programme. The statements made, and views expressed are solely the responsibility of the Fellow.

This research also was funded by the University of Rwanda through the SIDA open grant 2021-2023.

## Acknowledgement

I acknowledge everyone who supported and contributed to this study, including the participants and research assistants from the five referral hospitals, especially Joel Nshumuyiki, the chief research assistant. Special thanks go to my Supervisors for the PhD project entitled” Health Outcomes of long term disabilities following Road Traffic Injuries in Rwanda”.

## Data availability statement

Data supporting the study findings are available on request from the corresponding author [JAI]. The data are not publicly available due to ethical data transfer restrictions of IRB that could compromise the privacy of research participants.

## Disclaimer

The views and opinions expressed in the submitted article are the author’s own and not the official position of the affiliated institutions.

## Contributions

- JAI, JBS, AS, CU, DM, DT, GU, and GB participated in all stages of this paper, from the study design, methodology, grant writing, data collection, analysis and paper writing.

